# Seroprevalence of SARS-CoV-2 significantly varies with age: preliminary results from a mass population screening

**DOI:** 10.1101/2020.06.24.20138875

**Authors:** Gabriele Pagani, Federico Conti, Andrea Giacomelli, Dario Bernacchia, Rossana Rondanin, Andrea Prina, Vittore Scolari, Cecilia Eugenia Gandolfi, Silvana Castaldi, Giuseppe Marano, Cosimo Ottomano, Patrizia Boracchi, Elia Biganzoli, Massimo Galli

**Author notes:** Equally contributing Authors (Elia Biganzoli, Massimo Galli). **Corresponding author:**, Gabriele Pagani, Infectious Diseases department, III Division, ASST FBF-Sacco, Via G.B. Grassi 74, 20157, Milano, 02 3904 259s1.

## Abstract

**Objectives:** Castiglione D’Adda is one of the municipalities more precociously and severely affected by the Severe Acute Respiratory Syndrome Coronavirus 2 (SARS-CoV-2) epidemic in Lombardy. With our study we aimed to understand the diffusion of the infection by mass seroprevalence screening.

**Methods:** We searched for SARS-CoV-2 IgGs in the entire voluntary population using lateral flow immune-cromatographic tests on capillary blood (rapid tests). We then performed chemioluminescent serological assays (CLIA) and naso-pharyngeal swabs in a randomized representative sample of 562 subjects and in every subject with a positive rapid test.

**Results:** Based on CLIA serologies on the representative random sample (509 subjects), we estimated a 23% IgG seroprevalence. We also found a strong correlation between age and prevalence, with the elderly showing the highest probability of a positive serological test.

**Conclusions:** In an area of unrestricted viral circulation less than one-fourth of the population tested positive for SARS-CoV-2 IgG. Seroprevalence increased with increasing age, possibly suggesting differences in susceptibility to the infection.

## Introduction

Coronavirus Induced Disease 2019 (COVID-19) is caused by a novel *betacoronavirus* which was first identified in China and denominated SARS-CoV-2 (Severe Acute Respiratory Sindrome Coronavirus 2) [1]. Italy was the first European country that suffered a wide spread of the infection, which caused hundreds of thousands of cases [2].

The municipality of Castiglione d’Adda, a rural town of about 4550 inhabitants located South-East of Milan, has been heavily affected by SARS-CoV-2 infection since the earliest stages of the epidemic and subjected to movement restrictions since February 23^rd^, 2020. As of June 21, 2020, 184 confirmed cases of COVID-19 were reported, the large majority of which requiring hospitalization, accounting for about 4% of the total population. At the same time, 47 deaths were officially attributed to COVID-19. During the epidemic, testing was restricted to severely symptomatic cases. Consequently, the true extent of the SARS-CoV-2 infection remains unknown.

## Methods

In this study, the entire population of Castiglione D’Adda was invited to perform a lateral-flow immunocromatographic tests on capillary blood (Prima Lab, Switzerland) from the 18^th^ of May to the 7^th^ of June. News about the mass screening was disseminated by the town municipality. A random sample of 562 subjects (stratified per sex and age) was invited to undergo confirmatory tests by chemiluminescent method on venipuncture drawn blood (CLIA, IgG anti-SARS-CoV-2, Abbott, USA) and SARS-CoV-2 PCR on NPS, regardless of RICT results. More detailed information about the randomization procedure and the study design are available on the complete protocol, published on MedXriv pre-print server [3].

The analysis of IgG prevalence in the different age groups was performed by logistic regression models with response variable equal to 1 for positive IgG results, and 0 for negative IgG results. Age and gender were included as independent variables. Results were reported in terms of estimated probabilities of being positive to IgG test as a function of age, with respective 95% confidence intervals.

## Results

Results presented in this paper are based on 509 people selected in the random sample who agreed to undergo venipuncture to perform CLIA serologies. Characteristics of the selected population are reported in Table 1A.

**Table 1A:** Characteristics of 509 subjects in the random sample. Numerical variables are presented as means. CAD: Coronary Artery Disease; MI: Miocardial Infarction; COPD: Chronic Obstructive Pulmonary Disease;

|  | **IgG negative**<br>**(n=394)** | **IgG positive**<br>**(n=115)** |
| --- | --- | --- |
| Gender (Female) | 200 (50.8%) | 49 (42.6%) |
| Age (years) | 46.0, 20.6 | 55.4, 19.5 |
| Contact with verified case | 93 (23.6%) | 61 (53.0%) |
| Smoker | 92 (23.4%) | 10 (8.7%) |
| Cardiovascular diseases |  |  |
| - CAD/MI | 10 (2.5%) | 3 (4.3%) |
| - Arrhythmias | 14 (3.6%) | 5 (4.3%) |
| - Hypertension | 68 (17.3%) | 32 (27.8%) |
| - Other | 14 (3.6%) | 14 (12.2%) |
| At least one of the above: | 84 (21.3%) | 47 (40.9%) |
| Rheumatic diseases | 19 (4.8%) | 11 (9.6%) |
| Diabetes mellitus | 12 (3.0%) | 6 (6.2%) |
| Chronic Lung diseases |  |  |
| - Asthma | 20 (5.1%) | 2 (1.7%) |
| - COPD | 1 (0.3%) | 1 (0.9%) |
| - Other | 9 (2.3%) | 4 (3.5%) |
| At least one of the above: | 29 (7.4%) | 7 (6.1%) |
| Oncological pathologies |  |  |
| Solid Tumors | 20 (5.1%) | 6 (5.2%) |
| Oncochematological | 2 (0.5%) | 2 (1.7%) |
| At least one of the above: | 22 (5.6%) | 8 (7.0%) |
| Symptomatic |  |  |
| - Fever | 65 (16.5%) | 66 (57.4%) |
| - Cough | 57 (14.5%) | 31 (27.0%) |
| - Anosmia | 23 (5.8%) | 37 (32.2%) |
| - Dysgeusia | 27 (6.9%) | 46 (40.0%) |
| - Dispnea | 23 (5.8%) | 13 (11.3%) |
| - Rush: | 11 (2.8%) | 4 (3.5%) |
| - Arthromyalgia | 34 (8.6%) | 36 (31.3%) |
| At least one of the above: | 124 (31.5%) | 89 (77.4%) |
| Other symptoms | 54 (13.7%) | 23 (20.0%) |

The overall seroprevalence found in the tested sample was 22.6% (95% confidence interval 17.2%- 29.1%). Interestingly, seroprevalence increases with increasing age (Table 1B).

**Tab.1B.** IgG positivity per age group.

| **Age**<br>**(years)** | **Number** | **IgG positive** |
| --- | --- | --- |
| 0-19 | 61 | 5 (8.19%) |
| 20-39 | 111 | 19 (17.1%) |
| 40-59 | 182 | 40 (22.0%) |
| >=60 | 155 | 51(32.9%) |
| **TOTAL** | **509** | **115 (22.6%)** |

In multivariate analyses, a significant effect of age was found (p<0.0001) while no significant association emerged between IgG positivity and gender (p=0.2560). The possible existence of a non-linear effect of age was tested by including spline polynomials, without significant results (p=0.9078). Furthermore, an age/gender interaction effect did not result significant (p=0.5199). Estimates of probabilities of being positive to IgG test, from a model including only age as independent variable, are reported in Fig.1.

**Fig.1:**
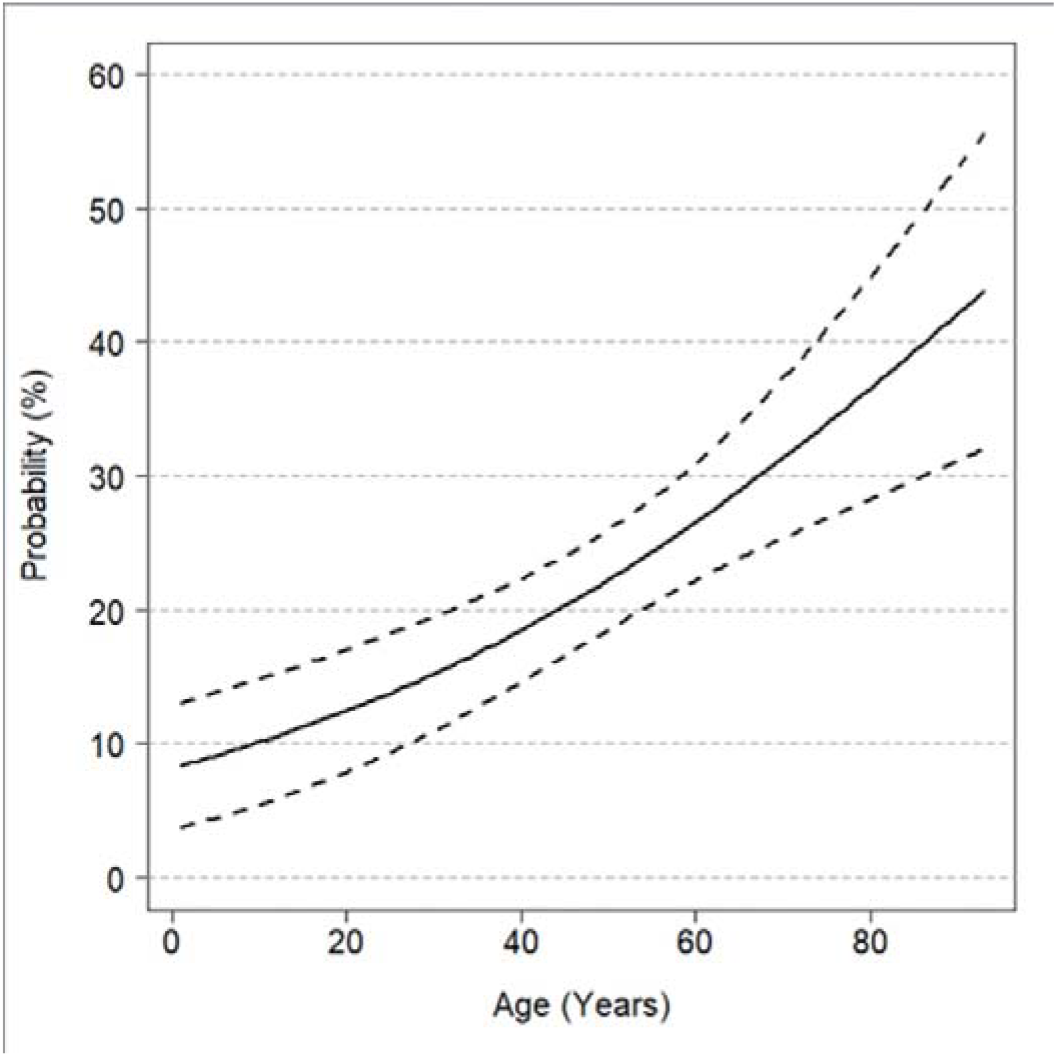
Estimated probability of IgG positivity as a function of age. Solid line: estimates, dashed lines: 95% confidence intervals.

## Discussion

Since the early phases of the pandemic, advanced age was identified as an independent predictor for severe disease and worse outcomes [4]. Beside this, it remains unclear if the limited number of cases reported in children [5] is due to a milder course of disease, with a larger percentage of asymptomatic cases, or to a lower susceptibility to infection, as our results seem to suggest. Different ACE2 expression according to age have been postulated to explain clinical expression and susceptibility to the infection. In particular, a higher expression of ACE2 in lung tissues in advanced age groups had been speculated [6, 7]. Moreover, a variable susceptibility to other coronavirus such as HCoV-NL63, which also use ACE2 as cell receptor in humans, in different age groups, has been also reported in different age groups [8].

Another possible explanation may be that an asymptomatic/pauci-symptomatic infection, more common in younger subjects, could elicit a less marked, or transient, antibody response, as already found in the closely related Middle East Respiratory Syndrome Coronavirus (MERS-CoV) [9]. A possible confounding factor in our findings could be related to social distancing measures: schools of any grade were among the first institutions to be closed in Italy, starting from the 5^th^ of March. This could have led to a lower exposure to the infection in children in pre-scholar and scholar age groups.

In conclusion, our findings suggest that IgG seroprevalence for SARS-CoV-2 increases with increasing age and these data suggest a lower susceptibility to infection in the lower age groups. These findings have important implications in epidemiology and public health, particularly in designing future population screenings, and could be an important contribution in the re-opening process, especially considered that more than three-fourths of the population could be still susceptible to SARS-CoV-2 infection, even in an area of initially unrestricted viral circulation.

## Data Availability

Complete dataset is available on a reasonable request.

## Acknowledgments

MG, GP, FC, DB, AG, RR and EB defined the study protocol. RR, AP, FC, DB and GP cooperated in the practical execution of the study and data gathering. GP drafted a first version of the manuscript, which was then revised and integrated by MG, EB, AG, CEG and SC. CO was responsible of serologies and NPSs execution. EB, PB and GM analyzed the data. All authors approved the final version of the manuscript.

## Ethical approval

The study was approved by University of Milan’s Ethical Committee.

